# Prevalence and molecular identification of *Schistosoma haematobium* infection in two peri-urban areas of Lusaka, Zambia: a cross-sectional study

**DOI:** 10.64898/2026.02.09.26345887

**Authors:** Mable Mwale Mutengo, James Mwansa, Kelly Chisanga, Emmanuel Zulu, Namwiinga Rozaria Mulunda, James Muchinga, Esther Rodríguez, Sergio Sánchez, Lourdes Castro, María Jesús Perteguer, David Carmena, Javier Sotillo

**Author notes:** Correspondence: Mable Mwale Mutengo. Institute of Basic and Biomedical Sciences, Levy Mwanawasa Medical University, Lusaka, Zambia., Javier Sotillo. Parasitology Reference and Research Laboratory, Centro Nacional de Microbiologia, Instituto de Salud Carlos III, Majadahonda, Madrid, Spain.

## Abstract

**Background:** Schistosomiasis is one of the most prevalent neglected tropical diseases in Sub-Saharan Africa, causing substantial morbidity and millions of disability-adjusted life years (DALYs). Although the WHO aims to eliminate schistosomiasis as a public health problem in several countries by 2030, limited data on infection prevalence in Zambia has hindered effective Mass Drug Administration (MDA) coverage, contributing to the persistence and resurgence of the disease.

**Methods:** We assessed the prevalence of urogenital schistosomiasis in two peri-urban districts of Lusaka (Chongwe and Kafue). A total of 208 participants were enrolled, and infection status was determined using microscopy, serological assays, and molecular diagnostics.

**Results:** Prevalence was significantly higher in Chongwe than in Kafue, as detected by both microscopy and qPCR, demonstrating a strong association between infection and district location. No association was found between sex and infection in either district. However, a significant association between age and *Schistosoma haematobium* infection was observed across both sites. Molecular characterization of individual eggs revealed that *S. haematobium × S. curassoni* hybrids were the most prevalent species circulating in the study population.

**Conclusions:** Our findings reveal a high prevalence of urogenital schistosomiasis in at least two peri-urban areas of Lusaka, Zambia, indicating that transmission remains highly active and may be underestimated in national surveillance. Furthermore, the presence of hybrid species infecting humans highlights the need to consider livestock reservoirs when designing elimination strategies. These results provide updated information on the epidemiological situation of urogenital schistosomiasis in Zambia and will support planning and implementation within the WHO NTD agenda.

**AUTHOR SUMMARY:** Urogenital schistosomiasis remains a major public health concern in many parts of Sub-Saharan Africa, yet recent data from Zambia are limited, particularly in communities surrounding Lusaka. In this study, we assessed infection levels in two peri-urban districts, Chongwe and Kafue, using a combination of microscopy, serology, and molecular techniques to ensure accurate detection. We found substantial differences in prevalence between the two locations, with significantly higher infection rates in Chongwe, as well as a clear association between age and risk of infection. Molecular characterisation of parasite eggs revealed that hybrid *Schistosoma haematobium × S. curassoni* forms were the dominant circulating genotypes, indicating potential involvement of animal reservoirs in transmission. These findings demonstrate that transmission remains active and likely underestimated, and they highlight the importance of incorporating updated epidemiological data and potential zoonotic sources into future control and elimination strategies in Zambia.

## INTRODUCTION

Schistosomiasis is a chronic, often debilitating disease, caused by various trematodes of the genus *Schistosoma* [1]. It is estimated that, worldwide, more than 250 million people are infected with *Schistosoma* spp., 90% of whom live in Africa [2]. Furthermore, these parasites cause tens to hundreds of thousands of direct deaths annually [3], while near 800 million people live in high-risk areas and over 250 million people required preventive chemotherapy in 2021 [2,4]. The Global Burden of Disease Study 2016 estimates that schistosomiasis accounts for 1.9 million disability-adjusted life years (DALYs) worldwide [5], although a previous meta-analysis study suggested a severalfold higher burden [6]. Of the six species that can infect humans, *Schistosoma haematobium* causes over half of all infections, leading to urogenital schistosomiasis and various health issues, including urinary and genital disorders [1,2]. Notably, urogenital schistosomiasis has been linked to squamous cell carcinoma of the urinary bladder. Indeed, the WHO International Agency for Research on Cancer (IARC) classifies *S. haematobium* as a Group 1 carcinogen [7,8].

Infection with *S. haematobium* occurs when the cercariae released from the snail intermediate host penetrate the skin of the human host, losing their tail and transforming into schistosomula [1]. After entering the venous blood vessels, the schistosomula are transported to the lungs, then to the heart, reaching the arterial circulation and finally migrating to the venous plexuses around the urinary bladder. Once there, females will produce and shed thousands of eggs daily, which pass through the bladder wall, and are excreted in the urine. Some eggs are trapped in the bladder, ureters, and genital tract contributing to host immune reactions. These eggs will hatch upon contact with water, releasing miracidia that penetrate the intermediate host snail (usually snails of the genus *Bulinus* in the case of *S. haematobium*).

Although both intestinal and urinary schistosomiasis are endemic in all 10 provinces of Zambia, the most prevalent species is *S. haematobium* [9]. Higher prevalences are usually found in rural districts near rivers and lakes [10], although schistosomiasis is also prevalent in many peri-urban areas due to poor water supply infrastructure [11]. For instance, the overall prevalence of schistosomiasis in children between ages of 5 and 17 years in a peri-urban area outside Lusaka was 20.7% [11], while in other areas from the Siavonga and Lusaka districts, the prevalence ranged between 8–12% [12]. However, most epidemiological studies have employed microscopy as the reference standard for diagnosis, which might have led to underdiagnosis of the disease, and prevalences could be higher [9,12]. A recently published meta-analysis showed that prevalence of schistosomiasis by *S. haematobium* declined between 1990–2010, most likely due to diagnostic and treatment efforts from the Zambian Bilharzia Control Programme (ZBCP), but has significantly increased in the last decade, which was attributed to a lack of monitoring and implementation of surveillance programs [9,13]. Despite some initiatives to conduct schistosomiasis surveys in Zambia in the early 2000’s, there is little information available on the prevalence of the infection in recent years. This has led to inefficient coverage of Mass Drug Administration (MDA) interventions and increase of other diseases such as HIV, which has been associated to schistosome infection in urban and peri-urban areas of Lusaka [14].

In the current study, we have performed an epidemiological survey in two different peri-urban areas of Lusaka: Chongwe and Kafue. These areas were selected based on previous mapping data which showed high schistosomiasis prevalence (>50% and near 90% prevalence in Chongwe and Kafue, respectively) [15]. To minimise the limitations of microscopy as a reference method (e.g. low sensibility) patients were screened for urinary schistosomiasis using molecular and serological methods in addition to a microscopy analysis. The results obtained herein provide important information about the current prevalence of schistosomiasis in two peri-urban areas of Zambia and can be of great value to design effective targeted MDA interventions against urogenital schistosomiasis.

## METHODS

### Ethics statement

The research was approved by the Levy Mwanawasa Medical University Research Ethics Committee (LMMU-REC 000054/23) and the authority to conduct research was obtained from the National Health Research Authority (NHRA0001/23/04/2023). Prior to commencement of the study, permission was also sought from the Lusaka Province Health office and the Ministry of Education. Informed consent was obtained from the parents or legal guardians of all participating children, and assent was obtained from the children themselves when appropriate. Participation was entirely voluntary, and care was taken to ensure that children and their guardians understood that they could withdraw from the study at any time without any consequence. The confidentiality and privacy of all participants were rigorously protected. Names of participants who were found with schistosomiasis were submitted to the Ministry of Health Neglected Tropical Diseases Unit for further action.

### Study area and population

The study was conducted in two different peri-urban regions of Lusaka Province namely Chongwe and Kafue Districts (Fig.1). Chongwe District is located 40 kilometres east of Lusaka Province, the capital city of Zambia. The district has a population of 313,389 inhabitants with most of its residents engaged in farming and livestock rearing. Chongwe District is traversed by the Chongwe River, which is a key water source for both domestic use and agriculture for most households. However, the river and its tributaries also pose a risk for waterborne diseases such as schistosomiasis.

**Fig 1.**
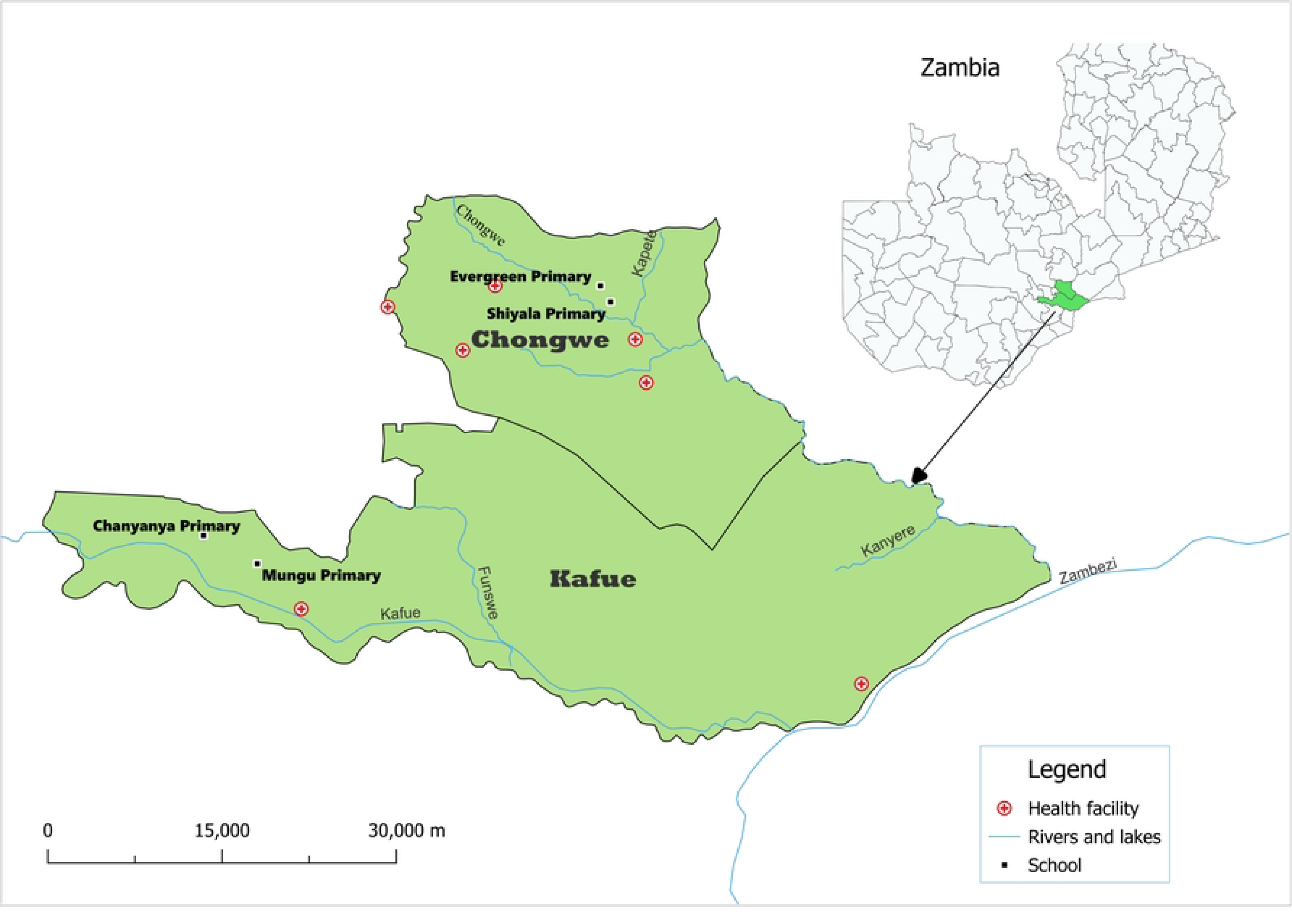
Map of the Chongwe and Kafue Districts in Lusaka Province showing the location of the sampled primary schools.

Kafue District is situated in the Southern part of Lusaka Province at the tip of Lusaka District and is located 45 kilometres away from the capital city, Lusaka. It has a population of 220,000 inhabitants. The district shares borders with Chongwe District in Northeast Lusaka Urban in the North, the Southwestern part borders Mazabuka and Chirundu districts. Kafue district has two major rivers, the Kafue and the Zambezi River running through the district with a sizable number of perennial rivers. This abundance of water creates several pockets of recreational water bodies where children go to swim and later are infected with bilharzia. Most of the district population is settled along the Zambezi and Kafue River where agriculture and fishing are important livelihood activities. Participants were school-age children who were selected using systematic random sampling method. They were recruited from two schools in Chongwe namely Evergreen and Shiyala basic schools and Kafue District namely Chanyanya and Mungu Basic Schools (Fig. 1).

### Parasitological and biochemical analysis

A single urine sample was collected from each of the recruited participants and analysed for visible haematuria, microscopic leucocytes, biochemistry protein parameters and presence of *S. haematobium* eggs. Visible haematuria and microhaematuria was detected macroscopically and with Haemastix^®^ dipsticks (Bayer, Germany) following the manufacturer’s instructions and recorded as negative, trace, 1+, 2+, 3+. For microscopy detection of *S. haematobium*, the urine filtration method was used. Briefly, each urine was vigorously mixed and 10 ml was drawn into a syringe and passed through a nucleopore membrane filter, with a pore size of 20 μm [16]. The filter was then placed onto a microscope slide and examined under low power (40×) magnification for the presence of *S. haematobium* eggs. The eggs were counted manually, and infection load was recorded as the number of eggs per 10 ml of urine.

### Recombinant protein production

The recombinant protein *Sh*-TSP-2 was expressed and purified as described previously [17]. In brief, *E. coli* BL21(DE3) harbouring the protein-encoding plasmid was cultured overnight at 37°C with agitation at 200 rpm in 10 ml of Luria broth supplemented with 50 μg/ml kanamycin (LB_kan_). Subsequently, the overnight culture was inoculated (1/100) into 500 ml of fresh LB_kan_ and grown at 37°C with shaking until reaching an OD_600_ of 0.5–1.0. Protein expression was induced by adding 1 mM isopropyl beta-D-1-thiogalactopyranoside (IPTG) and allowing the culture to incubate for an additional 24 h. After induction, cells were harvested by centrifugation at 8,000 *g* for 20 minutes at 4°C, then resuspended in 50 ml of lysis buffer (50 mM sodium phosphate pH 8, 40 mM imidazole, 300 mM NaCl), and subjected to three cycles of freeze/thawing followed by sonication at 4°C. The bacterial lysate was then centrifuged at 20,000 *g* for 20 min at 4°C, and the resulting supernatant was collected and stored at –80°C.

Recombinant proteins were purified using an ÄKTA Pure UPC FPLC system (GE Healthcare, IL, USA) equipped with Ni2+ immobilized metal ion affinity chromatography (IMAC) at 4°C. For this, the recombinant protein solutions were diluted 1:4 in lysis buffer, filtered through a 0.45 μm membrane, and loaded onto a 1 ml His-Trap IMAC column (GE Healthcare) pre-equilibrated with lysis buffer at a flow rate of 1 ml/min. Bound proteins were washed with 10 column volumes of lysis buffer, followed by elution with a linear gradient of imidazole (100–500 mM) in lysis buffer. Fractions containing the highest purity of the target protein were pooled and subjected to buffer exchange into PBS using an Amicon Ultra-15 centrifugal filter with a 3 kDa molecular weight cut-off. The identity of the purified proteins was confirmed through SDS-PAGE and Western blot analysis utilizing anti-His monoclonal antibodies (Invitrogen, CA, USA).

### ELISA assays

Nunc™ MicroWell™ 96-Well Medisorp™ Microplates (Thermo Fisher Scientific, MA, USA) were coated overnight at 4°C with a solution containing 2 μg/ml of *Sh*-TSP-2 in 0.1 M Na_2_CO_3_/NaHCO_3_ buffer at pH 9.6. The following day, the plates were washed with PBS-Tween-20 at 0.05% (PBST) and blocked for 2 h at room temperature with 100 μl of PBST/5% skimmed milk powder. Following blocking, 50 μl of urine (diluted 1:10 in PBST) were added to the respective wells and allowed to incubate overnight at 4°C. The following day, the plates were washed with PBST and subsequently incubated for 1 h at room temperature with 100 μl of goat anti-human IgG-HRP (Sigma, MA, USA) diluted to 1:5,000 in PBST. After another round of washing with PBST, the plates were developed using 3,3′,5,5′-Tetramethylbenzidine (TMB), and the reaction was halted with 0.5 M sulfuric acid. The absorbance was measured at a wavelength of 492 nm using a Multiskan FC (Thermo Fisher Scientific) microplate reader.

### DNA extraction and real-time PCR

DNA was extracted using the Quick-DNA Urine Kit (Zymo Research, CA, USA) following the manufacturer’s instructions. Briefly, 70 μL of Urine Conditioning Buffer was added for every 1 ml of urine, followed by the addition of 10 μl of Clearing Beads. The samples were, then, mixed and centrifuged at 3,000 *g* for 15 min. Subsequently, the Tough-to-Lyse Samples protocol was followed. The urine supernatant was decanted, leaving behind 200 μl of pellet. The pellet was transferred to a ZR BashingBead Lysis Tube (0.1 & 0.5 mm), and 600 μl of BashingBead Buffer was added. The mixture was vortexed in a Vortex-Genie^®^ 2 mixer for 5 min at maximum speed. Tubes were then centrifuged at 10,000 *g* for 1 min, and 400 μl of the sample was applied to a Zymo-Spin III-F Filter and centrifuged again at 10,000 *g* for 1 min. Following this, 1,200 μl of Genomic Lysis Buffer was added to the flow-through, and the mixture was centrifuged through a Zymo-Spin IC-S Column. The column was washed successively with 200 μl of Urine DNA Prep Buffer, and 700 μL and 200 μl of Urine DNA Wash Buffer. The columns were then transferred to clean 1.5 ml microcentrifuge tubes, and DNA was eluted by adding 50 µl of deionized molecular biology-grade water directly to the column matrix, followed by centrifugation at 14,000 *g* for 1 min. The concentration and purity of the extracted DNA were assessed using a Nanodrop One (Thermo Fisher Scientific) spectrophotometer.

Reactions targeting the Dra1 tandem repeat [18] were performed in duplicate as follows: 20 µl reactions contained 5 μl of DNA, 10 μl of Quantimix easy kit (Biotools, Madrid, Spain), 1 ul of PrimeTime^®^ standard qPCR Assay that included 500 nM of primers Fwd (5’–GATCTCACCTATCAGACGAA–3’) and Rv (5’–TCACAACGATACGACCAAC) each and 300 nM of probe (FAM-TGTTGGTGGAAGTGCCTGTTTCGCAA-IowaBlack Fluorescent Quencher). Extraction blanks and qPCR blanks were used as control for cut-off calculation. A Thermocycler (Rotor Gene Q, Qiagen, Germany) setting consisted of an initial step of 2 min at 95°C followed by 45 cycles of 15 s at 95°C and 60 s at 60°C.

### Molecular Characterisation of Parasite Eggs

A total of sixty-four *Schistosoma* individual eggs were carefully isolated form the urine samples of twenty-four (24) patients using a stereomicroscope and a pipette into prelabelled Eppendorf tubes. DNA was extracted using the Quick DNA urine kit (Zymo Research) as described by the manufacturers. The extracted DNA was stored at –20°C until analysis. A duplex Tetra-ARMS PCR assay which simultaneously genotypes the Internal Transcribed Spacer (ITS) and 18S ribosomal gene was performed to differentiate *S. haematobium*, *S. curassoni*, *Schistosoma bovis*, and their hybrids based on species-specific polymorphisms as previously described [19]. Briefly, the reaction mixture (20 µl total volume per reaction) consisted of 5 µl genomic DNA, 5 µl of 5× PCR buffer, 0.5 µl each of outer forward (OF-ITS, OF-18S) and outer reverse (OR-ITS, OR-18S) primers, 0.5 µl each of inner forward and reverse primers (IF-Sb/Sc-ITS, IF-Sb-18S, IR-Sh-ITS, IR-Sc/Sh-18S), 0.5 µl MyTaq DNA polymerase (Bioline, London, UK), and nuclease-free water to make up the final volume. Amplification was carried out in a GenAmp 2700 thermal cycler (Thermo Fisher Scientific) using the following conditions: an initial denaturation at 94 °C for 2 min; 35 cycles of denaturation at 94 °C for 1 min, annealing at 62 °C for 45 s, and extension at 72 °C for 40 s; followed by a final extension at 72 °C for 6 min. The reaction was held at 4°C until further analysis. PCR products were separated on a 1.8% agarose gel prepared in 0.5× TBE buffer. A volume of 3 µL of each amplicon was loaded, and electrophoresis was run at 135 V for 60 minutes. DNA bands were visualized under UV light following gel red staining. The ITS/18S T-ARMS-PCR provides unique amplicon profiles for each species as follows: *Sh*:120, 382, 487 and 667 bp; *Sc*: 120, 234, 382 and 667 bp; *Sb*: 234, 316, 382, and 667 bp, and/or hybrid forms (*Sb*×*Sc*: 120, 234, 316, 382 and 667 bp; *Sh*×*Sc*: 120, 234, 382, 487 and 667 bp; and *Sh*×*Sb*: 120, 234, 316, 382, 487 and 667 bp).

### Sequencing of the Nuclear Ribosomal Internal Transcribed Spacer (ITS) Gene

Twenty-nine DNA samples out of the 64 DNA samples extracted from individual eggs were subjected to Sanger sequencing of the ITS gene to characterise the *Schistosoma* species. Briefly, ITS bands observed either at 487 bp or at 234 bp were excised from the agarose gel and DNA extracted using the Isolate II PCR and Gel Kit (Meridian Bioscience, OH, USA). Samples were sequenced in both directions using ITS primers (outer and inner) described above by capillary electrophoresis using the BigDye Terminator chemistry (Applied Biosystems) on an on ABI PRISM 3130 automated DNA sequencer. Raw chromatograms were visually inspected for quality control and for detecting the presence of ambiguous (double peak) positions, edited, and assembled into consensus sequences using BioEdit v7.2.5 software. Consensus sequences were subjected to BLASTn searches against the NCBI nucleotide database to determine species identity and identify any potential hybrids. All sequences have been deposited in Genbank (accession numbers PX953088-PX953107).

#### Phylogenetic and Sequence Analysis

ITS consensus sequences were aligned using Muscle with default parameters (https://www.ebi.ac.uk/jdispatcher/msa/muscle). Reference sequences representing *S. bovis*, *S. curassoni*, *S. haematobium*, *S. haematobium* x *S. curassoni*, *S. haematobium* x *S. bovis* and *S. mansoni*, were retrieved from GenBank and included for comparison. *S. turkestanicum* sequences were also retrieved from GenBank and used as an outgroup to root the tree. A phylogenetic tree was constructed using IQ-TREE v1.6.12 [20]. The best-fit substitution model was automatically selected using ModelFinder. Branch support was assessed using ultrafast bootstrap approximation with 10,000 replicates, SH-like approximate likelihood ratio test with 1,000 replicates, and approximate Bayes test. Additional weighted bootstrap trees and nearest-neighbor interchange moves were included to improve tree search. The final consensus tree was computed from bootstrap replicates and written in NEXUS format.

### Statistical Analysis

The prevalence of infection and its 95% confidence interval (CI) were calculated using an in-house Python script, incorporating the Z-value for a 95% confidence level (https://github.com/JavierSotillo/Scripts/blob/main/Prevalence_CI_Calculator.py). All other statistical tests were performed using GraphPad Prism v 10.2. A two-proportion z-test (Chi-squared test) was employed to study the association between location, sex or age and *Schistosoma* infection. To calculate the correlation between age and the number of *Schistosoma* eggs in infected children, a Spearman correlation test was performed. To compare the mean age of male and female participants, as well as the median number of eggs in each location, a two-tailed Mann-Whitney test was employed. *P*-values < 0.05 were considered as significant.

## RESULTS

### 3.1 Demographic and Biochemical Characteristics of the Study Participants

A total of 208 participants from Chongwe and Kafue districts were enrolled into the study (111 from Chongwe and 97 from Kfaue) (Supplementary Table 1). From these, a total of 99 were females and 109 were males, and their mean age was 10.3 (95% CI = 9.7 to 10.9) and 10.83 (95% CI = 10.3 to 11.4), respectively (Table 1). No significant difference between the males and females in terms of age was found (*P* = 0.184). From the 111 participants enrolled in the Chongwe district, a total of 55 were females and 56 were males, and their mean age was 11.6 (95% CI = 10.8 to 12.4) and 11.9 (95% CI = 11.2 to 12.7), respectively (Table 1). Furthermore, from the 97 participants enrolled in the Kafue district, a total of 44 were females and 53 were males, and their mean age was 8.7 (95% CI = 8.1 to 9.3) and 9.6 (95% CI = 9.00 to 10.3), respectively (Table 1). Mean age differences were statistically significant in the case of Kafue district (*P* < 0,05).

Only 22 (10.6%) from the total participants showed visible haematuria, whereas microhaematuria was positive by Hemastix^®^ in 53 participants (25.5%). From these, 3 of them showed traces, and 16, 13 and 21 were categorized as 1+, 2+ and 3+ based on the band intensities (Supplementary Table1). Leukocytes were detected in the urine of 40 participants (19,23%), with intensities varying from trace (7 participants), 1+ (26 participants), 2+ (6 participants) and 3+ (1 participant). Only 29 of these leukocyte positive participants also showed microhaematuria (Supplementary Table1).

### 3.2 Significant Association Between *Schistosoma* Infection and District Location in Two Zambian Peri-Urban Districts

First, to determine the prevalence of *S. haematobium* infection in two different districts from Zambia (Chongwe and Kafue), a microscopy analysis was performed as this is the gold-standard diagnostic technique recommended by WHO. *Schistosoma haematobium* eggs were detected in the urine samples of 48 children, corresponding to an overall prevalence of 23.1% (95% CI = 17.3 to 28.8) and a median number of eggs in 10 ml of 54.5 (95% CI = 45.0 to 79.0) (Table 2). The Spearman correlation coefficient between age and the number of *Schistosoma* eggs in infected children was -0.016 (*P* = 0.904), indicating a weak non-significant negative correlation. From the 48 participants positive to eggs by microscopy, 34 (70.8%) belonged to the Chongwe district and 14 (29.2%) from Kafue, resulting in a significant association between *Schistosoma* positivity by microscopy and district location (χ^2^ = 4.84; *P* < 0.05) (Table 2). The median number of eggs found was 51.5 (95% CI = 35.0 to 79.0) in positive participants from Chongwe, while it was 71 (95% CI = 36.0 to 209.0) in positive participants from Kafue. No statistically significant differences were found when comparing the median number of eggs in each location.

Since sensitivity of microscopy is low compared to molecular methods, we conducted a prevalence analysis using qPCR targeting the Dra1 repetitive motif as described above. In this case, a total of 110 children were positive to infection, corresponding to a prevalence of 52.9% (95% CI = 46.1 to 59.7). Of these, 82 positive children were from Chongwe district, while 28 from Kafue. These results showed a prevalence of 73.9% (95% CI = 65.7 to 82.1) in Chongwe district and 28.9% (95% CI = 19.9 to 37.9) in Kafue district, indicating a significant association between *Schistosoma* positivity and district location (χ^2^= 13.52; *P* < 0.01) (Table 2).

Interestingly, no statistically significant association with geographic location was found when investigating the prevalence of schistosomiasis by ELISA (χ^2^ = 0,29; *P* > 0.5). A total of 48 children were positive by ELISA in both districts (46.2%), corresponding to a prevalence of 43.2% (95% CI = 34.0 to 52.5) in Chongwe District and 49.5% (95% CI = 39.5 to 59.4) in Kafue District (Table 2).

### 3.3 District-Level Differences in the Influence of Sex and Age on *Schistosoma* Infection

When analysing microscopy results by sex, out of the 48 schistosomiasis positive children, 31 (14.9%; 95% CI = 10.1–19.7) were males and 17 (8.2%; 95% CI = 4.5–11.9) were females. As expected, higher prevalences were observed among males (29.8%; 95% CI = 23.6–36.0) than females (22.6%; 95% CI = 16.9–28.3) by qPCR and ELISA (26.0%; 95% CI = 20.0–31.9 and 22.1%; 95% CI = 16.5–27.7). However, no significant association between *Schistosoma* positivity and sex was observed (Table 3).

Similar results were observed when analysing microscopy results by sex and location. Among the 111 participants in Chongwe, 34 were positive for *S. haematobium* eggs (20 males and 14 females) (Table 3). Interestingly, among the 97 participants from Kafue, 14 were positive by microscopy (11 males and only 3 females). However, no statistically significant difference in the proportion of *S. haematobium* egg-positive cases between males and females was observed in either Chongwe or Kafue (Table 3). Based on the microscopy results, the overall median egg account among all positive children was 54.5 (95% CI: 45.0–79.0) corresponding to a median of 56 (95% CI: 36–107) for males and of 49 (95% CI: 43.0 to 103.0) for females. No significant difference between the median number of eggs between males and females was observed (*P* = 0.7448).

Similarly, among the 82 qPCR-positive children from Chongwe (*n* = 111), 44 were males and 38 females, while in Kafue, 19 males (67.9%) and 9 (32.1%) females were positive out of 97 participants (Table 3). In the case of ELISA, similar prevalences were observed for males and females in both districts (22.5–23.4% and 21.7–28.9%, respectively). No significant association between *Schistosoma* infection and sex was observed within individual districts when prevalence was analysed by qPCR or ELISA.

A statistically significant association between age and *S. haematobium* infection was identified across both study locations (*P* < 0.0001). The median age of infected children was 12 years (95% CI: 11.0–12.0), compared to 10 years (95% CI: 9.0–10.0) among uninfected children. This trend was also evident in Kafue (p < 0.05), where the median age of infected children was 10 years (95% CI: 8.0–12.0), versus 9 years (95% CI: 8.0–10.0) for those uninfected. However, no significant age-related difference was observed in Chongwe.

### 3.4. Phylogenetic analysis revealed clear clustering patterns consistent with species identity and geographic origin

To determine the most prevalent *S. haematobium* forms (either pure or hybrid) in both districts, the eggs of twenty-four (24) participants were analysed using a duplex tetra-primer ARMS-PCR assay described previously [19] and ITS bands subjected to Sanger sequencing. From these, sequencing of 7 participants failed during the process. T-ARMS PCR and subsequent Sanger sequencing of positive bands revealed that infections were due either to infections with *S. haematobium* pure forms or with *S. haematobium* x *S. curassoni* hybrids.

For the phylogenetic analyses, *S. haematobium* ITS sequences from other locations as well as other *Schistosoma* species were included to obtain a broader picture of the evolutionary relationships and geographic distribution of hybridization events among *Schistosoma* species in Zambia. Individual sequences from both regions grouped within the *S. haematobium* clade, showing strong bootstrap support (>80%) for their placement alongside reference *S. haematobium* sequences from Africa (Figure 2). No sequences from Kafue or Chongwe clustered with *S. mansoni* or *S. turkestanicum*, confirming species specificity. Two samples, SC 32-5 (Chongwe) and 159-5 (Kafue), were positioned within the *S. haematobium* clade and in proximity to hybrid references (*S. haematobium × S. bovis*), although they did not form a distinct subcluster together. Additionally, all other sequences from Kafue and Chongwe clustered within the *S. haematobium* clade together with African reference isolates (e.g., Zimbabwe and Zambia), showing strong bootstrap support (>80%). This grouping reflects high ITS similarity and confirms species identity, with no evidence of separate geographic clades for the two Zambian regions.

**Fig. 2.**
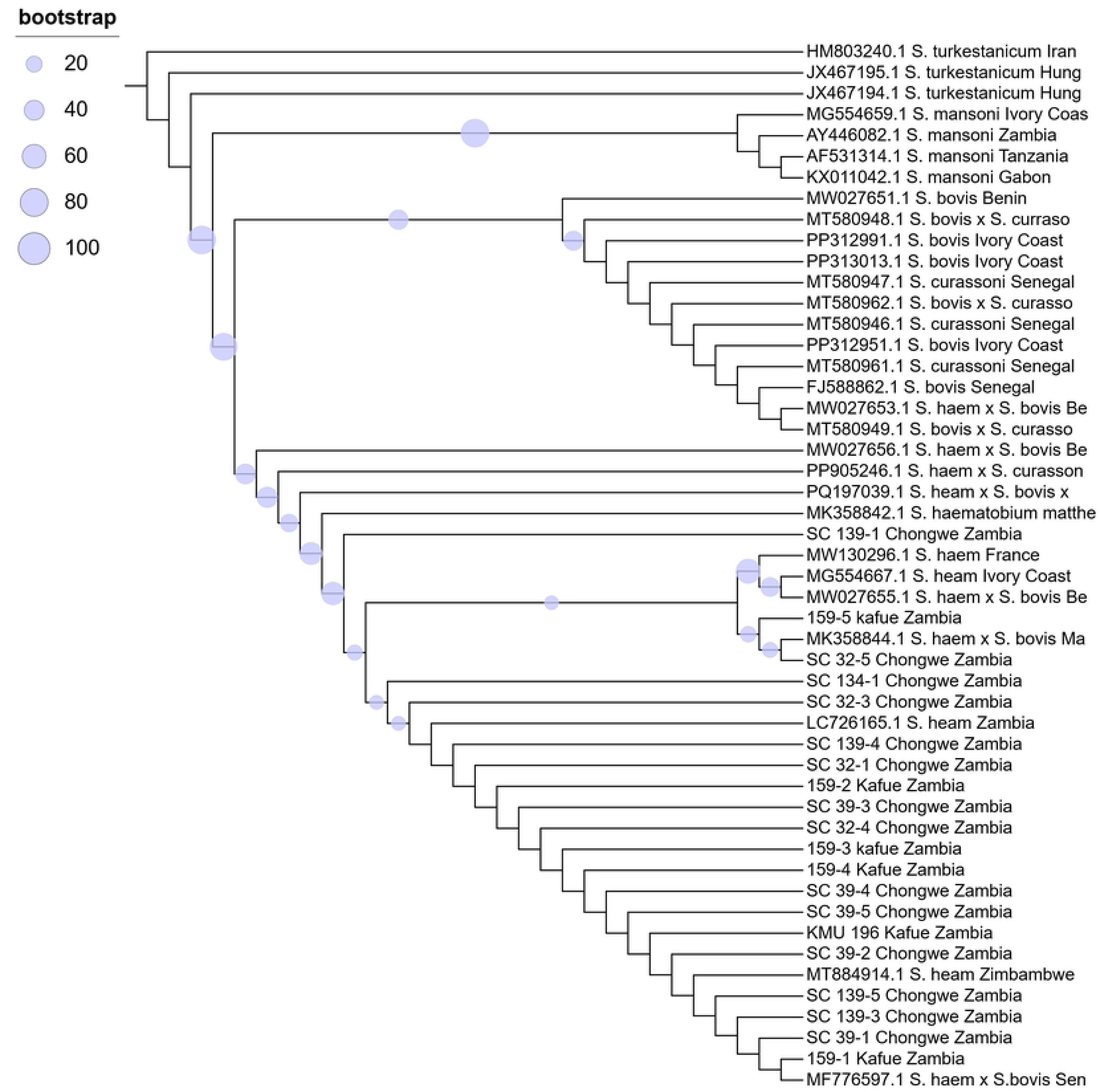
Phylogenetic tree based on ITS sequences from *Schistosoma* eggs collected in Kafue and Chongwe, Zambia.

## DISCUSSION

Schistosomiasis remains a significant public health concern in sub-Saharan Africa, with persistently high prevalence rates driven by systemic gaps in diagnostic capacity and access to curative interventions. In Zambia, this neglected tropical disease (NTD) affects up to 90% of the population in high-transmission provinces, disproportionately impacting children, women, and impoverished rural communities. Among women, female genital schistosomiasis is particularly concerning, with an estimated 20–56 million affected across Africa. This condition is associated with infertility, adverse pregnancy outcomes, and a threefold increased risk of HIV acquisition [21]. Beyond its physical health consequences, schistosomiasis undermines social and economic wellbeing, especially in underserved regions where it is recognized as a driver of poverty and cyclical re-poverty [22]. In this context, obtaining accurate, region-specific prevalence data for *S. haematobium* infections in Zambia is critical for informing the strategic deployment of MDA programs and progressing toward the 2030 targets for NTD elimination [23]. To contribute to this effort, we assessed the prevalence of urogenital schistosomiasis in two peri-urban districts of Lusaka Province (Chongwe and Kafue) using different diagnostic approaches. Additionally, we investigated the presence of hybrid *Schistosoma* species in these regions through a duplex tetra-ARMS PCR assay followed by sequencing in order to confirm species identity and detect potential introgressive hybridization events that may influence transmission dynamics and control strategies.

Overall, the qPCR approach exhibited higher sensitivity (52.9%; 95% CI: 46.1–59.7) compared to conventional microscopy (23.1%; 95% CI: 17.4–28.8). This aligns with previous studies, as qPCR is widely recognized for its superior diagnostic performance over microscopy, particularly due to its ability to detect low-intensity infections and trace amounts of parasite DNA that may be missed by traditional parasitological methods [24,25]. In contrast, our serological findings revealed slight lower sensitivity for schistosomiasis diagnosis relative to qPCR (46.2%; 95% CI: 39.1–52.9). Furthermore, a higher rate of false positives was observed in our ELISA results. This phenomenon has been well documented, as residual antibodies can persist in serum and urine for several months after praziquantel treatment, potentially leading to an overestimation of active infections, particularly, though not exclusively, in endemic areas [26–28]. Interestingly, the majority of false positives identified in this study were detected mainly in the Kafue district. Although there is no documented evidence of previous MDA campaigns in the area, the elevated number of false positives may be due to recent praziquantel treatments administered to children in the region, either for schistosomiasis or for other helminth infections. Such treatments may have contributed to a reduction in disease burden, but not necessarily to the persistence of antibodies. As a result, serological findings in this region may not accurately reflect current active infections.

Interestingly, a significant association was observed between age and infection status among the studied children. Overall, the median age of infected children was 12 years, compared to 10 years among uninfected children. In Kafue specifically, median ages were 10 and 9 years for infected and uninfected participants, respectively. The influence of age on *Schistosoma* infection has been well documented [29–31]. For example, Lamberti *et al*. reported that *Schistosoma* spp. infections peaked at ages 11–12 in Uganda [31], while a meta-analysis on *S. mansoni* risk factors identified the 13–16 age group as highest risk, followed by 10–12 years [30]. This trend likely reflects the age at which children engage more in activities such as swimming, increasing their exposure to *S. haematobium* compared to younger children. Additionally, older participants may have developed partial acquired immunity, which influences infection dynamics and shapes the age–intensity curves observed for *S. haematobium* [32].

Both microscopy and qPCR analyses also revealed a statistically significant association between geographic location and the prevalence of urogenital schistosomiasis. Prevalence rates were markedly higher in Chongwe compared to Kafue (70.8% vs. 29.2% by microscopy; 73.9% vs. 28.9% by qPCR), with differences reaching statistical significance (*P* < 0.05). These findings suggest that the observed disparity in infection rates between the two districts is unlikely due to chance and instead reflects distinct schistosomiasis risk profiles. In Zambia, schistosomiasis is typically more prevalent in rural areas located near water bodies such as rivers and lakes [10]. However, peri-urban regions are also affected, largely due to inadequate water infrastructure [11]. For instance, in a peri-urban community near Lusaka, the infection rate among children aged 5 to 17 years was reported at 20.7% [11], while other locations within the Siavonga and Lusaka districts showed prevalence rates ranging from 8% to 12% [12]. In Chongwe, exposure is primarily linked to agricultural and domestic use of the Chongwe River. In contrast, in Kafue, recreational and fishing activities around the Kafue and Zambezi Rivers contribute significantly to transmission risk, particularly among children. Previous studies investigating protozoan parasite prevalence in these regions also found higher rates of the diarrhoea-causing enteric protozoan *Cryptosporidium parvum* among schoolchildren in Chongwe compared to Kafue [33]. This was attributed to increased exposure to livestock and zoonotic transmission in Chongwe, where cattle and small ruminant farming is a key economic activity [33].

Notably, more than half of the participants screened for parasite genotyping were found to be infected with a hybrid *Schistosoma* species, predominantly *S. haematobium × S. curassoni*. While previous studies in Zambia have reported hybrid forms involving other livestock schistosomes, such as *S. haematobium × S. bovis* and *S. mattheei × S. haematobium* [12,34], this is, to our knowledge, the first documented occurrence of *S. curassoni* hybrids in the country. This novel finding suggests active zoonotic transmission cycles and underscores the potential role of livestock as reservoir hosts, which may complicate control and elimination efforts. Given that *S. curassoni* is typically associated with livestock in West Africa, its detection in Zambian human populations may indicate either the introduction of infected animals or underrecognized local transmission involving animal reservoirs [35]. Furthermore, this observation aligns with the higher prevalence of urogenital schistosomiasis in Chongwe compared to Kafue observed in our study and reinforces the significance of livestock in sustaining transmission.

The tight clustering of Kafue and Chongwe sequences with African *S. haematobium* references indicates limited genetic differentiation across these sites, consistent with high gene flow within endemic areas. However, the proximity of certain samples, such as SC 32-5 (Chongwe) and 159-5 (Kafue), to hybrid references (*S. haematobium × S. bovis*) suggests possible introgression events. Hybridization between human and animal schistosome species can expand host ranges, creating zoonotic reservoirs that sustain transmission even when human treatment coverage is high. This complicates elimination strategies that rely solely on MDA in humans, as untreated livestock may act as persistent sources of infection, and may facilitate the spread of schistosomiasis [36]. Indeed, the predominance of *S. haematobium × S. curassoni* hybrids also underscores the need for integrated One Health approaches that consider both human and animal health in endemic regions [37]. It is important to note, however, that the T-ARMS PCR assay employed in this study was specifically designed to detect *S. haematobium*, *S. curassoni*, and *S. bovis*. As such, the presence of other hybrid combinations, potentially involving *S. mattheei* or additional *Schistosoma* species, cannot be excluded. Future studies employing broader molecular tools, such as whole-genome sequencing or high-resolution genotyping, will be essential to fully characterize the diversity and epidemiological significance of hybrid *Schistosoma* species in Zambia.

This study presents several limitations that should be taken into account when interpreting the findings and drawing conclusions. Firstly, the survey was restricted to paediatric populations in only two districts of Lusaka Province, which may limit the generalizability of both the prevalence and molecular data to the broader Zambian context. Secondly, as a cross-sectional study, it did not allow for longitudinal tracking of infections in individual children or assessment of seasonal trends in pathogen occurrence. Thirdly, since participation was voluntary, there is a possibility of selection bias, families who perceived a higher risk of infection may have been more inclined to take part, potentially skewing the results, especially in areas with lower response rates. Lastly, the low number of patients screened for the genotyping of the parasites may have hampered the identification of other potential hybrid parasites in the samples.

In conclusion, we have performed the most up-to-date prevalence study of urogenital schistosomiasis in the paediatric population in two regions of the Lusaka Province, Zambia. We have demonstrated the presence of *S. haematobium × S. curassoni* hybrids in these two regions of Zambia, probably related to direct contact with small ruminants as part of the local economic activities. Our results underscore the need for integrated One Health approaches, combining human and veterinary health interventions, to effectively reduce transmission and prevent the establishment and spread of hybrid schistosome populations in endemic communities.

## Data Availability

All sequences generated in this study have been deposited in Genbank (accession numbers PX953088-PX953107)
https://www.ncbi.nlm.nih.gov/nuccore/PX953088
https://www.ncbi.nlm.nih.gov/nuccore/PX953089
https://www.ncbi.nlm.nih.gov/nuccore/PX953090
https://www.ncbi.nlm.nih.gov/nuccore/PX953091
https://www.ncbi.nlm.nih.gov/nuccore/PX953092
https://www.ncbi.nlm.nih.gov/nuccore/PX953093
https://www.ncbi.nlm.nih.gov/nuccore/PX953094
https://www.ncbi.nlm.nih.gov/nuccore/PX953095
https://www.ncbi.nlm.nih.gov/nuccore/PX953096
https://www.ncbi.nlm.nih.gov/nuccore/PX953097
https://www.ncbi.nlm.nih.gov/nuccore/PX953098
https://www.ncbi.nlm.nih.gov/nuccore/PX953099
https://www.ncbi.nlm.nih.gov/nuccore/PX953100
https://www.ncbi.nlm.nih.gov/nuccore/PX953101
https://www.ncbi.nlm.nih.gov/nuccore/PX953102
https://www.ncbi.nlm.nih.gov/nuccore/PX953103
https://www.ncbi.nlm.nih.gov/nuccore/PX953104
https://www.ncbi.nlm.nih.gov/nuccore/PX953105
https://www.ncbi.nlm.nih.gov/nuccore/PX953106
https://www.ncbi.nlm.nih.gov/nuccore/PX953107

## SUPPORTING INFORMATION

**Supplementary Table 1.** Spreadsheet summarizing demographic information and diagnostic indicators for study participants, including district, age, sex, urine dipstick results, *Schistosoma haematobium* egg counts, and test results by microscopy, qPCR, and ELISA.

**Supplementary Figure 1.** Representative image of a duplex Tetra-ARMS PCR assay performed on individual schistosome eggs. Lanes 1-5: samples. B: blank control. *Sh* x *Sc* +: Positive control for *Schistosoma haematobium* x *Schistosoma curassoni* hybrid. *Sb*+: Positive control for *Schistosoma bovis*.

